# PROGNOSTIC VALUE AND RELATIVE CUT-OFFS OF TRIGLYCERIDES PREDICTING CARDIOVASCULAR OUTCOME IN A LARGE REGIONAL-BASED ITALIAN DATABASE

**DOI:** 10.1101/2023.06.23.23291840

**Authors:** Valérie Tikhonoff, Edoardo Casiglia, Agostino Virdis, Guido Grassi, Fabio Angeli, Marcello Arca, Carlo M. Barbagallo, Michele Bombelli, Federica Cappelli, Rosario Cianci, Arrigo FG Cicero, Massimo Cirillo, Pietro Cirillo, Raffaella Dell’oro, Lanfranco D’elia, Giovambattista Desideri, Claudio Ferri, Ferruccio Galletti, Loreto Gesualdo, Cristina Giannattasio, Guido Iaccarino, Francesca Mallamaci, Alessandro Maloberti, Stefano Masi, Maria Masulli, Alberto Mazza, Alessandro Mengozzi, Maria Lorenza Muiesan, Pietro Nazzaro, Paolo Palatini, Gianfranco Parati, Roberto Pontremoli, Fosca Quarti-Trevano, Marcello Rattazzi, Gianpaolo Reboldi, Giulia Rivasi, Elisa Russo, Massimo Salvetti, Pier Luigi Temporelli, Giuliano Tocci, Andrea Ungar, Paolo Verdecchia, Francesca Viazzi, Massimo Volpe, Claudio Borghi

## Abstract

**Background:** Despite longstanding epidemiologic data on the association between increased serum triglycerides (TG) and cardiovascular (CV) events, the exact level at which risk begins to rise is unclear. The Working Group on Uric Acid and Cardiovascular Risk of the Italian Society of Hypertension has conceived a protocol aimed at searching for the prognostic cut-off value of TG in predicting CV events in a large regional-based Italian cohort.

**Methods:** Among 14,189 subjects aged 18 to 95 years followed-up for 11.2 (5.3-13.2) years, by means of receiver operating characteristic (ROC) curve the prognostic cut-off value of TG, able to discriminate combined CV events, was identified. The conventional (150 mg/dL) and the prognostic cut-off values of TG were used as independent predictors in separate multivariate Cox models adjusted for age, sex, body mass index, total and high-density lipoprotein cholesterol, serum uric acid, arterial hypertension, diabetes, chronic renal disease, smoking habit, use of antihypertensive and lipid lowering drugs.

**Results:** During 139,375 person-years of follow-up, 1601 participants experienced CV events. ROC curve showed that 89 mg/dL (95%CI 75.8-103.3, sensitivity 76.6, specificity 34.1, p<0.0001) was the prognostic cut-off value for CV events. Both cut-off values of TG, the conventional and the newly identified, were accepted as multivariate predictors in separate Cox analyses, the hazard ratios being 1.211 (95%CI 1.063-1.378, p=0.004) and 1.150 (95%CI 1.021-1.295, p=0.02), respectively.

**Conclusions:** Lower (89 mg/dL) than conventional (150 mg/dL) prognostic cut-off value of TG for CV events do exist and it is associated with increased CV risk in an Italian cohort.

**Clinical Perspective:**

1. **What is new?**

1. Evidence indicates that elevated triglyceride levels are related to cardiovascular events and mortality. However, the exact level at which risk begins to increase is unclear.
2. In a large cohort of European subjects, a prognostic cut-off value of triglycerides lower (89 mg/dL) than the conventional one (150 mg/dL) was identified.
2. **What are the clinical implications?**

1. Triglyceride measurement must be considered an important part of the routine evaluation to manage cardiovascular risk.
2. In primary prevention, subjects with triglycerides above 89 mg/dL should be carefully observed to prevent possible cardiovascular events.

## Introduction

The global burden of dyslipidemia has increased over the past 30 years, with elevated plasma low-density lipoprotein (LDL) cholesterol levels being the eighth most important risk factor for death in 2019 [1]. In high-income countries (mostly in Europe and North America), levels of LDL have been in steady decline in response to improvements in lifestyle and an increase in the use of lipid-lowering drugs [2,3]. Based on risk stratification, LDL cholesterol lowering is addressed aggressively by statin and by nonstatin therapies (ezetimibe, inhibitors of proprotein convertase subtilisin/kexin type 9, bempedoic acid, evinacumab, and inclisiran) today [4,5]. Despite reduction on cardiovascular (CV) mortality over the past two decades, the number of deaths remains high and a residual risk persists [6]. This has compelled an interest in individuals with residually elevated triglyceride (TG), who have higher concentrations of atherogenic cholesterol carried by circulating TG-rich lipoproteins [7–10]. It became more and more clear that if LDL is well controlled and non high-density lipoprotein (HDL) cholesterol is not well controlled, the culprit is TG-rich lipoprotein which is highly atherogenic [9,10].

The interest on the association between elevated TG and CV events has fluctuated over the past many years driven by changes in the evidence base that suggests that these either cause atherosclerotic CV disease or simply represent innocent bystanders [8]. Mendelian randomization studies have provided causal evidence for the role of TG-mediated pathways in coronary heart disease (CHD) incidence [11]. In the large meta-analysis from Emerging Risk Factors Collaboration study, comprising 302,430 people without an initial vascular disease compiled from 68 long-term prospective studies of Europe and North America, a total of 12,785 cases of CHD were recorded from a total of 2.79 million person-years of follow-up, showing a hazard ratio for nonfatal myocardial infarction and CHD death for TG as 1.37 (95% CI, 1.31–1.42) after adjustment for non-lipid risk factors [12]. In a recent paper, Raposeiras-Roubin et al. shown in a prospective cohort study including 3,754 middle-aged individuals with low to moderate CV risk that TG levels ≥150 mg/dl was associated with subclinical atherosclerosis and vascular inflammation, even in participants with normal LDL-C levels [13].

Despite epidemiologic data demonstrating the association between elevations in serum TG and CV disease [14], the exact level at which risk begins to increase is unclear. The first lipid guidelines for CV prevention defined elevated TG as >250 mg/dL [15]. Since then clinical trials evaluating the impact of TG lowering therapies have used a cutoff around 150 mg/dL [16]. Therefore, in the recent European Society of Cardiology (ESC) guidelines, the treatment target and goal on CV prevention indicates the conventional cut-off of 150 mg/dl [4]. However, specific prognostic cut-off value of TG around which the rise of incident CV events associated with TG changes appears has not been precisely evaluated in a European cohort. The Working Group on Uric Acid and Cardiovascular Risk of the Italian Society of Hypertension has conceived and designed an ad hoc protocol aimed at searching for prognostic cut-off values of TG in predicting CV events in a large regional-based Italian cohort of men and women.

## Methods

### Database and study protocol

The database called URRAH involves data on subjects aged 18 to 95 years collected on a regional community basis from all the territory of Italy with a median follow-up period of 11.2 years (interquartile range from 5.3-13.2 years) up to 31 July 2017. The study protocol has been previously extensively described [17–19] and the STROBE cohort checklist was used to write the article [20]. The data that support the findings of this study are available from the corresponding author upon reasonable request. In brief, a nationwide Italian database was built by collecting data on subjects from representative cohorts having serum uric acid measurement and complete information about several variables including outcomes. 14,189 subjects were taken into account in the present analyses. For all subjects, a standardized set of items was recorded, including demographics, anthropometric measures, metabolic parameters, smoking habit, systolic and diastolic arterial blood pressure, renal function, history of CV, renal and brain disease, concomitant treatments and outcomes. Hypertension was defined by the presence of at least two blood pressure recordings ≥140 or ≥90 mmHg or treatment with antihypertensive medications. Diabetes mellitus was defined if blood glucose was ≥126 mg/dl at fast or ≥200 mg/dl 2 hours after 75 g oral glucose load or if glycated haemoglobin was ≥48 mmol/mol. Kidney function was evaluated through estimation of the glomerular filtration rate, using a standardized serum creatinine assay and according to the Chronic Kidney Disease Epidemiology Collaboration equation [21]. Chronic kidney disease (CKD) was defined for estimated glomerular filtration rate values <60 mL/min per 1.73 m^2^. Procedures for taking and preparing blood specimens and laboratory analysis were standardized. Blood specimens were taken by venipuncture after an overnight fast. Specimens were placed in edetic acid tubes in an ice bath. Plasma was then separated in a refrigerated centrifuge at 4 °C within 2 h after collection; separated plasma was transferred into cryovials, and frozen for later measurement of lipid concentration. The study data were collected routinely or ad hoc in previously authorized studies. Subjects underwent no extra tests or interventions, and there was no impact on subjects’ care or outcome.

### Ethics

The study data were collected routinely or *ad hoc* in previously authorized studies. Subjects underwent no extra tests or interventions, and there was no impact on subjects’ care or outcome. The study was performed according to the Declaration of Helsinki for Human Research (41^st^ World Medical Assembly, 1990). The processing of the patients’ personal data collected in this study comply with the European Directive on the Privacy of Data. All data to be collected, stored and processed are anonymized, and all study-related documents are retained in a secure location. No personal information is stored on local personal computers. Approval was sought from the Ethical Committee of the Coordinating Center at the Division of Internal Medicine of the University of Bologna (No. 77/2018/Oss/AOUBo). Informed consent was obtained from all subjects at recruitment.

### Outcome

According to the study protocol [18], incident events due to acute myocardial infarction, *angina pectoris*, heart failure, stroke, transient ischemic attack and hypertensive complications were taken into consideration during the follow-up (see Table 1s in Supplemental Materials for ICD10 codes). Events were double-checked with hospital and physicians’ files.

### Statistics

#### General description

The SAS package version 9.4 (SAS Institute, Cary, NC) was used for statistical analysis. A preliminary power analysis based on differences from stratified values of TG for α=0.05 and power (1-β)=0.80 was performed. Power analysis showed that the number of subjects in the database (n=14,189) represented a sample largely sufficient to avoid β error. The Kolmogorov-Smirnov normality test was performed. Continuous variables were expressed as mean ± standard deviation and compared among classes or categories by the analysis of covariance adjusted time to time for proper confounders and followed by the Bonferroni’s post hoc test. Categorical variables were compared by means of the Pearson χ^2^ test. In multivariate analyses, the covariables that were not independent from each other were previously log-transformed. The null hypothesis was rejected for values of p<0.05.

#### Preliminary Cox analysis

Multivariate dichotomic Cox regression models having all combined CV events (fatal + non-fatal) as dichotomic dependent variable, adjusted for age, sex, body mass index, serum uric acid, serum HDL-cholesterol, serum non-HDL cholesterol, arterial hypertension, diabetes, chronic renal disease, smoking habit, alcohol consumption and use of lipid lowering drugs were preliminarily used to search for an association between TG log-transformed as a continuous variable and CV event. We tested interactions of TG log-transformed with sex, diabetes mellitus, arterial hypertension, ethanol intake by incorporating corresponding interaction terms in the analysis. Hazard ratios (HR) with 95% confidence intervals (CI) were produced. The null hypothesis was rejected for values of p<0.05.

#### Univariate prognostic cut-off values

The receiver operating characteristic (ROC) curves method was used to search for prognostic cut-off of TG for CV events in the whole database. TG was used as basic variable and CV events as dichotomic classification variable. The De Long et al. method [22] was used. Ratio of cases in the positive group (prevalence), sensitivity and specificity were calculated. ROC curve was generated in the whole database, and a prognostic cut-off value was identified as the curve point nearest to the 100% of axis of the ordinates [23]. In practical terms, this was made by identifying the TG value associated to the highest values of the sum sensitivity + specificity. Youden’s index [24] defined for all points of ROC curves was used as a criterion for selecting the optimum cut-off. The cut-off point identified is the value corresponding with maximum of the Youden index J=max[Se_i_ + SP_i_ -1], where Se_i_ and SP_i_ are the sensitivity and specificity over all possible threshold values. The area under the curve was also shown for each ROC curves analysis [25].

#### Validation of the conventional and prognostic cut-off values and HR of being over cut-off

The conventional (≥150 mg/dl) and the prognostic (identified by mean of the ROC curve) cut-off values of TG were used as independent variables in separate multivariate Cox analyses adjusted for the confounders already identified, having combined CV events as dichotomic dependent variable in the whole database. A cut-off value identified *via* the ROC curves method was considered as valid if accepted in the model being the null hypothesis rejected, otherwise it was considered a false cut-off. The corresponding HR with 95% CI were obtained. The conventional (≥150 mg/dl) and prognostic validated cut-off values were used in the whole database to stratify combined CV events in descriptive analysis and for generating outcome curves according to the Kaplan-Meier non-parametric estimator of limit product. Log-rank tests were used to assess differences between curves.

## Results

### Descriptive statistics

The general characteristics of the 14,189 subjects are shown in Table 1, also showing men and women separately. Very few participants (3.8%) were on lipid-lowering therapy. TG was non-normally distributed in the whole database with a median TG level of 110 (25^th^-75^th^ percentile 80-154) (Figure 1).

**Table 1.**
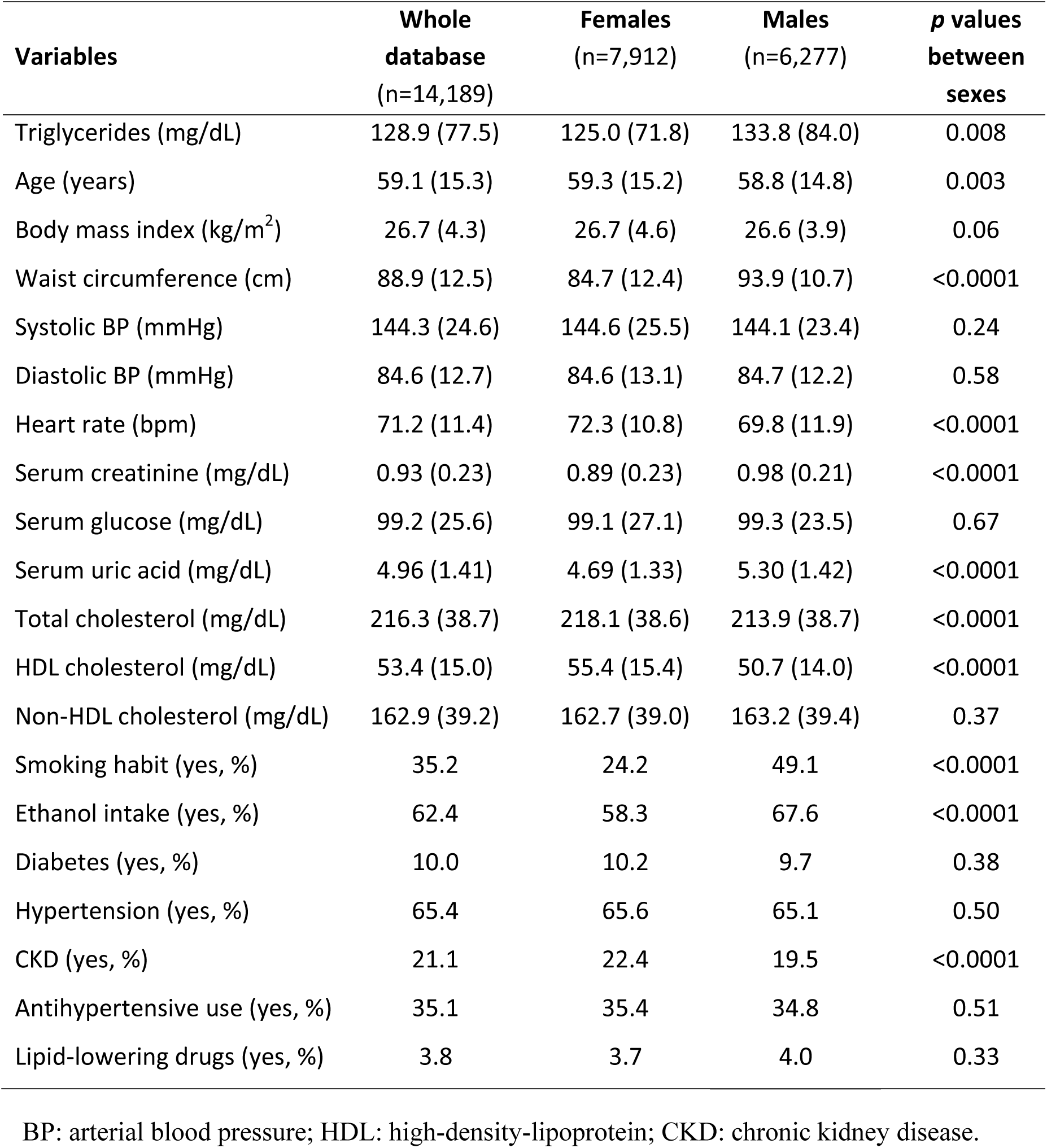
General characteristics of the study participants also showing sex stratification. Continuous variables are expressed as mean (standard deviation). Categorical variables are in %.

| **Variables** | **Whole**<br>**database**<br>**(n=14,189)** | **Females**<br>**(n=7,912)** | **Males**<br>**(n=6,277)** | **p values**<br>**between**<br>**sexes** |
| --- | --- | --- | --- | --- |
| Triglycerides (mg/dL) | 128.9 (77.5) | 125.0 (71.8) | 133.8 (84.0) | 0.008 |
| Age (years) | 59.1 (15.3) | 59.3 (15.2) | 58.8 (14.8) | 0.003 |
| Body mass index (kg/m<sup>2</sup>) | 26.7 (4.3) | 26.7 (4.6) | 26.6 (3.9) | 0.06 |
| Waist circumference (cm) | 88.9 (12.5) | 84.7 (12.4) | 93.9 (10.7) | <0.0001 |
| Systolic BP (mmHg) | 144.3 (24.6) | 144.6 (25.5) | 144.1 (23.4) | 0.24 |
| Diastolic BP (mmHg) | 84.6 (12.7) | 84.6 (13.1) | 84.7 (12.2) | 0.58 |
| Heart rate (bpm) | 71.2 (11.4) | 72.3 (10.8) | 69.8 (11.9) | <0.0001 |
| Serum creatinine (mg/dL) | 0.93 (0.23) | 0.89 (0.23) | 0.98 (0.21) | <0.0001 |
| Serum glucose (mg/dL) | 99.2 (25.6) | 99.1 (27.1) | 99.3 (23.5) | 0.67 |
| Serum uric acid (mg/dL) | 4.96 (1.41) | 4.69 (1.33) | 5.30 (1.42) | <0.0001 |
| Total cholesterol (mg/dL) | 216.3 (38.7) | 218.1 (38.6) | 213.9 (38.7) | <0.0001 |
| HDL cholesterol (mg/dL) | 53.4 (15.0) | 55.4 (15.4) | 50.7 (14.0) | <0.0001 |
| Non-HDL cholesterol (mg/dL) | 162.9 (39.2) | 162.7 (39.0) | 163.2 (39.4) | 0.37 |
| Smoking habit (yes, %) | 35.2 | 24.2 | 49.1 | <0.0001 |
| Ethanol intake (yes, %) | 62.4 | 58.3 | 67.6 | <0.0001 |
| Diabetes (yes, %) | 10.0 | 10.2 | 9.7 | 0.38 |
| Hypertension (yes, %) | 65.4 | 65.6 | 65.1 | 0.50 |
| CKD (yes, %) | 21.1 | 22.4 | 19.5 | <0.0001 |
| Antihypertensive use (yes, %) | 35.1 | 35.4 | 34.8 | 0.51 |
| Lipid-lowering drugs (yes, %) | 3.8 | 3.7 | 4.0 | 0.33 |

BP: arterial blood pressure; HDL: high-density-lipoprotein; CKD: chronic kidney disease.

**Figure 1.**
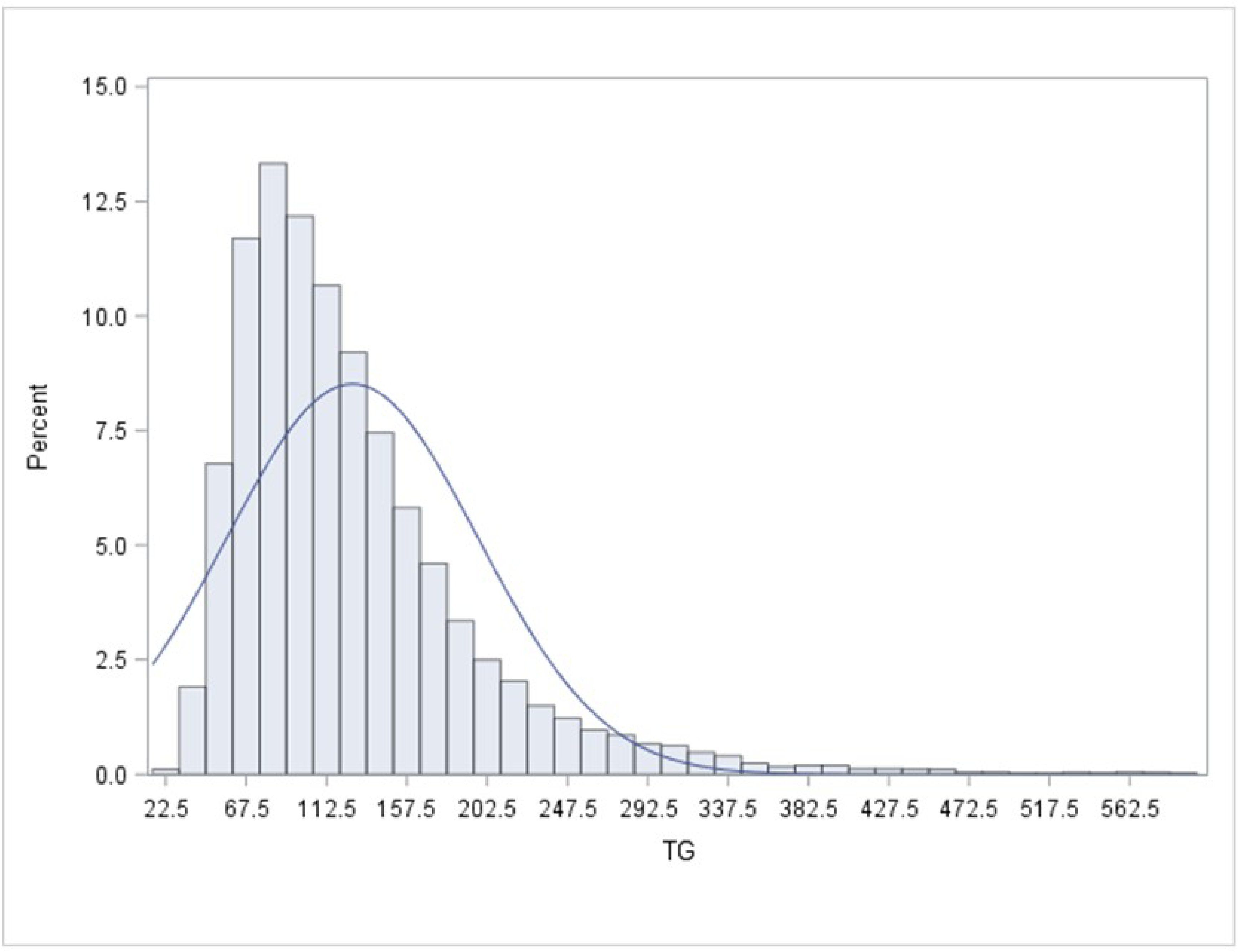
Distribution of baseline serum triglycerides in the whole database. TG: serum triglycerides.

During 139,375 person-years of follow-up, 1601 participants experienced CV events (11.4 per 1000 age-adjusted person-years), 747 men (12.1 per 1000 age-adjusted person-years) and 854 women (11.0 per 1000 age-adjusted person-years).

#### Multivariate analysis

Preliminary Cox models having combined CBV events as dependent variable showed that, in the whole cohort, log transformed TG as a continuous variable was a significant predictor of CV events [HR 1.173 (1.035-1.330), p=0.01] with significant confounding factors age, BMI, serum HDL cholesterol, serum uric acid, diabetes mellitus, arterial hypertension, alcohol consumption and use of lipid lowering drugs (Table 2). Four interaction terms were tested (TG x gender, TG x diabetes mellitus, TG x arterial hypertension, TG x ethanol intake). Only TG x diabetes mellitus was significant when included in the model (p=0.017) with HR 1.329, 95% CI 1.051–1.680.

**Table 2.** Cox model for combined cardiovascular events using serum triglycerides as a continuous independent variable in the whole cohort (n=14,189).

| Independent variables | HR | 95% CI | *p* value |
| --- | --- | --- | --- |
| Serum triglycerides (mg/dL) | 1.173 | 1.035-1.330 | 0.01 |
| Age (years) | 1.061 | 1.056-1.067 | <0.0001 |
| Sex (1=men, 0=women) | 1.025 | 0.919-1.144 | 0.66 |
| Body mass index (kg/m<sup>2</sup>) | 0.970 | 0.957-0.982 | <0.0001 |
| HDL cholesterol (mg/dl) | 0.992 | 0.989-0.996 | <0.0001 |
| Non-HDLcholesterol (mg/dL) | 0.999 | 0.998-1.001 | 0.44 |
| Serum uric acid (mg/dL) | 2.047 | 1.681-2.492 | <0.0001 |
| Diabetes (1=yes, 0=no) | 1.908 | 1.689-2.156 | <0.0001 |
| Hypertension (1=yes, 0=no) | 1.469 | 1.281-1.684 | <0.0001 |
| CKD (1=yes, 0=no) | 0.910 | 0.808-1.025 | 0.12 |
| Smoking (1=yes, 0=no) | 1.051 | 0.936-1.180 | 0.40 |
| Ethanol (1=yes, 0=no) | 1.380 | 1.216-1.566 | <0.0001 |
| Lipid-lowering drugs (1=yes, 0=no) | 0.621 | 0.477-0.809 | <0.001 |

HR: hazard ratios; CI: confidence intervals; HDL high-density-lipoprotein; CKD: chronic kidney disease.

#### Search for prognostic cut-off value of TG

ROC curve furnished plausible univariate cut-off value of TG (>88 mg/dL, 95%CI 75.8-103.3, sensitivity 76.6, specificity 34.1, p<0.0001) as prognostic cut-off value for CV event (Figure 2 and Table 3). When the conventional TG value of 150 mg/dL is considered, the sensitivity and specificity parameters are 33.0 and 74.3, respectively.

**Figure 2.**
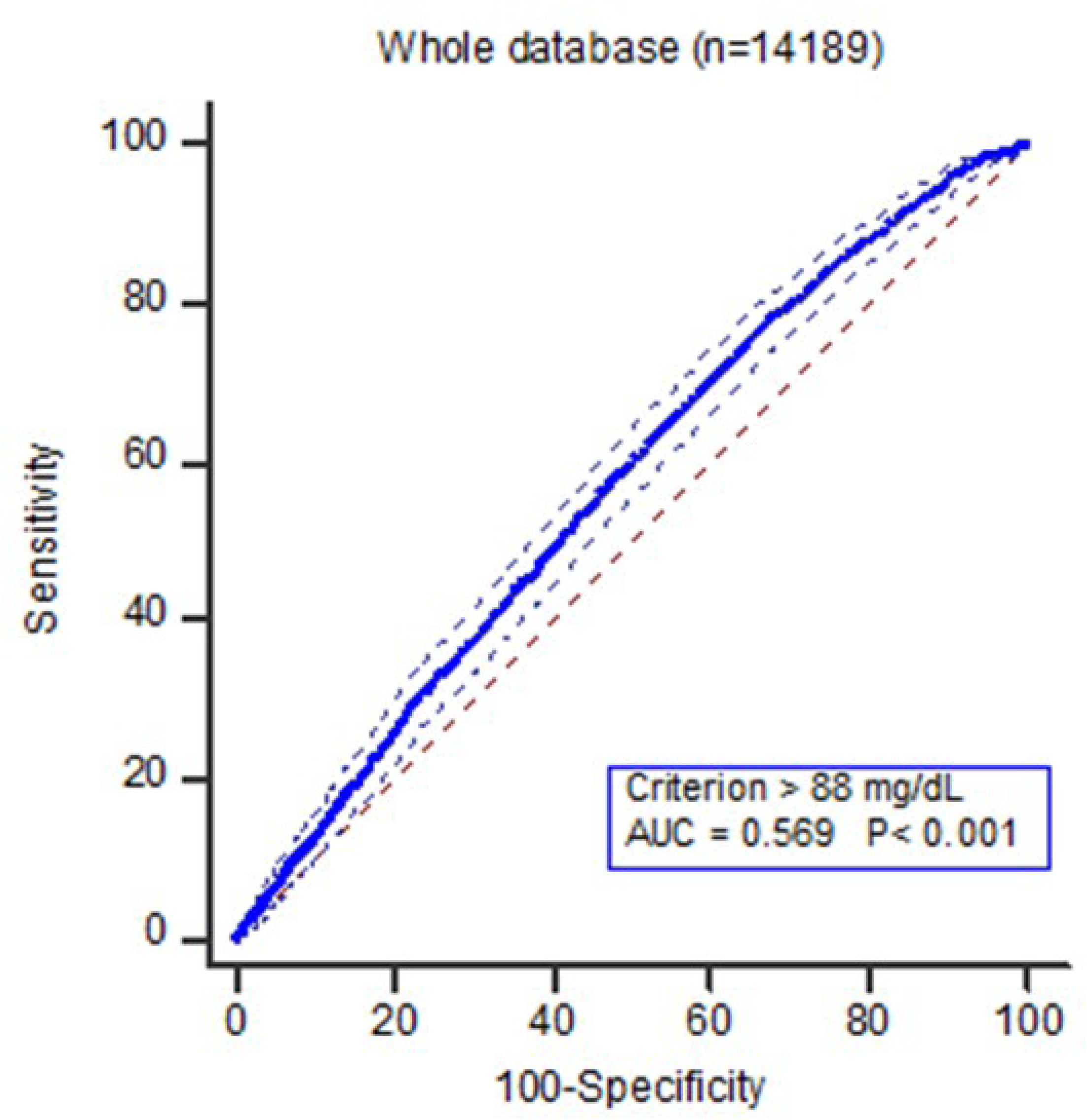
Receiver-operator-characteristic (ROC) curves of combined cardiovascular events. 95% confidential intervals are shown (thin lines). AUC: area under the curve; p: criterion for rejection of the null hypothesis.

**Table 3.** ROC curve parameters of the cut-off value for combined cardiovascular events in a regional community-based cohort of 14,189 subjects.

|  | All (n=14,189) |
| --- | --- |
| Cut-off (CI<sup>‡</sup>) | >89 mg/dL (75-103) |
| AUC (SE, CI) | 0.569 (0.007, 0.561-0.578) |
| Youden index (CI) | 0.1064 (0.0772-0.1233) |
| Sensitivity% (CI) | 76.6 (74.4-78.6) |
| Specificity% (CI) | 34.1 (33.2-34.9) |
| Z statistics, *p* | 9.521, <0.0001 |

CI: 95% confidence intervals; AUC: area under the curve;
SE: standard error; <sup>‡</sup>bootstrap confidence intervals (1000 iterations)

#### Validation of conventional and prognostic cut-off values and HRs of being over cut-off

The conventional (≥150 mg/dl) and the identified by the ROC curve (≥89 mg/dl) cut-off values of TG were both accepted as predictors in separate multivariate Cox analyses, adjusted for the confounders already identified, demonstrating that being over the cut-off values lead to higher risk with the 1.150 (95%CI 1.021-1.295, p=0.02) and HR 1.211 (95%CI 1.063-1.378, p=0.004), respectively (Table 4 and 5). Kaplan-Meier curves after stratification according to cut-off values of TG are shown in Figure 3. The curves of subjects having TG≤cut-off and TG>cut-off were clearly separate both for conventional (150 mg/dl) and prognostic (89 mg/dl) cut-off values.

**Table 4.** Hazard ratios of the variable “*over cut-off value of serum TG (≥150 mg/dL)”* for combined cardiovascular events in the whole database.

| Independent variables | HR | 95% CI | p value |
| --- | --- | --- | --- |
| Specific cut-off of TG ($\geq 150$ mg/dL) | 1.150 | 1.021-1.295 | 0.02 |
| Age (years) | 1.062 | 1.057-1.068 | <0.0001 |
| Sex (1=men, 0=women) | 1.028 | 0.921-1.146 | 0.63 |
| Body mass index (kg/m<sup>2</sup>) | 0.968 | 0.956-0.981 | <0.0001 |
| HDL cholesterol (mg/dL) | 0.992 | 0.989-0.996 | <0.0001 |
| Non-HDL cholesterol (mg/dL) | 0.999 | 0.998-1.001 | 0.72 |
| Serum uric acid (mg/dL) | 2.005 | 1.650-2.436 | <0.0001 |
| Diabetes (1=yes, 0=no) | 1.935 | 1.727-2.183 | <0.0001 |
| Hypertension (1=yes, 0=no) | 1.317 | 1.141-1.523 | 0.0002 |
| CKD (1=yes, 0=no) | 0.838 | 0.740-0.949 | 0.005 |
| Smoking (1=yes, 0=no) | 1.047 | 0.936-1.175 | 0.44 |
| Ethanol (1=yes, 0=no) | 1.444 | 1.270-1.638 | <0.0001 |
| Statin use (1=yes, 0=no) | 0.598 | 0.458-0.779 | <0.0001 |

HR: hazard ratios; CI: confidence intervals; TG: serum triglycerides; HDL: high-density-lipoprotein; CKD: chronic kidney disease;

**Table 5.** Hazard ratios of the variable “*over cut-off value of serum TG (≥89 mg/dL)”* for combined cardiovascular events in the whole database.

| Independent variables | HR | 95% CI | p value |
| --- | --- | --- | --- |
| Specific cut-off of TG ($\geq 89$ mg/dL) | 1.211 | 1.063-1.378 | 0.004 |
| Age (years) | 1.062 | 1.057-1.068 | <0.0001 |
| Sex (1=men, 0=women) | 1.028 | 0.921-1.146 | 0.62 |
| Body mass index (kg/m<sup>2</sup>) | 0.968 | 0.956-0.981 | <0.0001 |
| HDL cholesterol (mg/dL) | 0.992 | 0.989-0.996 | <0.0001 |
| Non-HDL cholesterol (mg/dL) | 0.999 | 0.998-1.001 | 0.54 |
| Serum uric acid (mg/dL) | 1.993 | 1.639-2.423 | <0.0001 |
| Diabetes (1=yes, 0=no) | 1.942 | 1.719-2.193 | <0.0001 |
| Hypertension (1=yes, 0=no) | 1.318 | 1.141-1.523 | 0.0002 |
| CKD (1=yes, 0=no) | 0.838 | 0.740-0.949 | 0.005 |
| Smoking (1=yes, 0=no) | 1.051 | 0.936-1.180 | 0.40 |
| Ethanol (1=yes, 0=no) | 1.442 | 1.270-1.638 | <0.0001 |
| Statin use (1=yes, 0=no) | 0.595 | 0.456-0.777 | <0.0001 |

HR: hazard ratios; CI: confidence intervals; TG: serum triglycerides; HDL: high-density-lipoprotein; CKD: chronic kidney disease;

**Figure 3.**
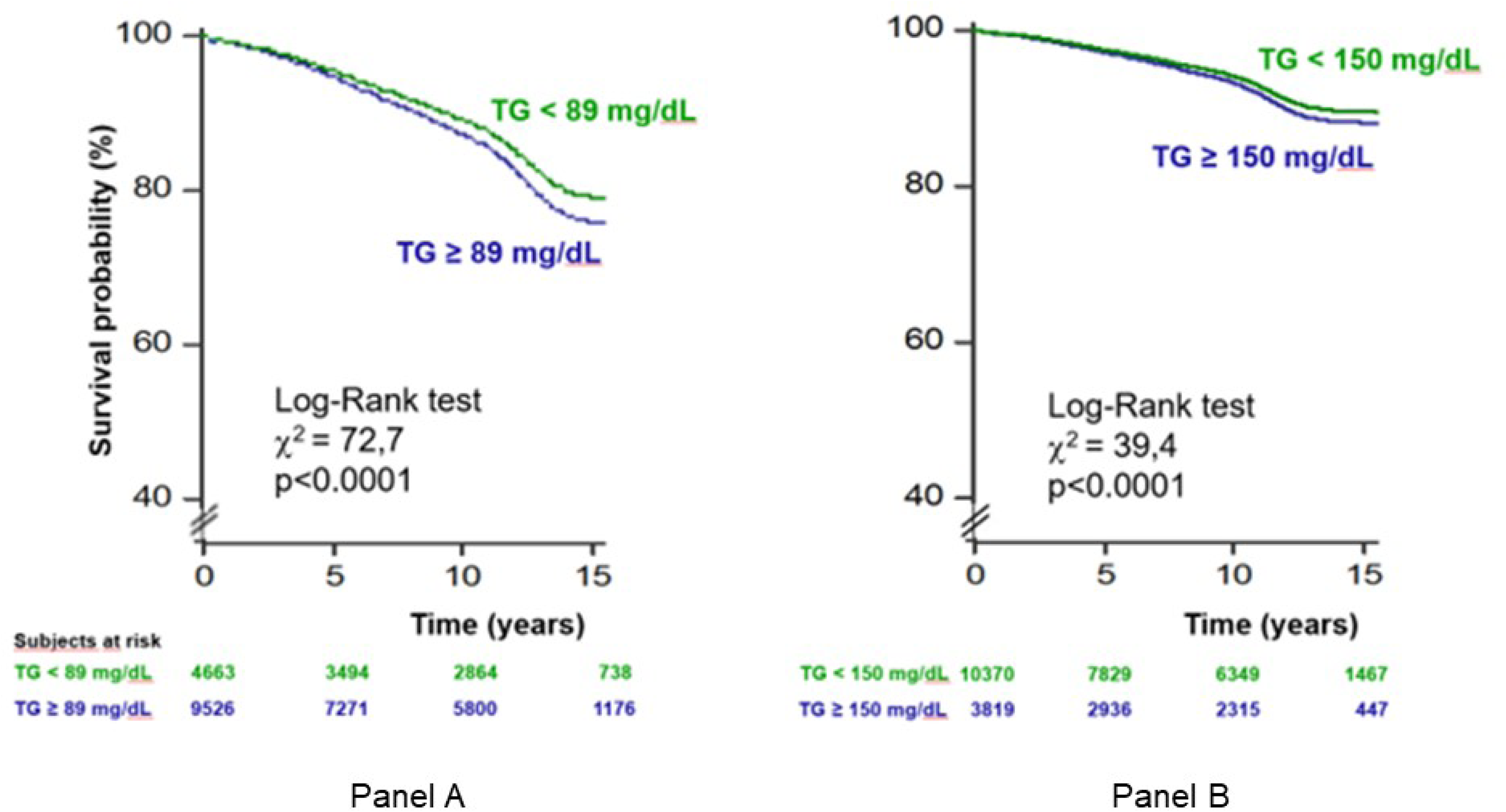
Kaplan-Maier survival curves for combined cardiovascular events for the identified cut-off value of triglycerides 89 mg/dL (panel A) and the conventional cut-off value of triglycerides 150 mg/dL (panel B). Trends of subjects having serum triglycerides > cut-off (blue line) and < cut-off (red line) are shown. Numbers of subjects at risk are shown in the two footnotes. Values of p indicate statistical difference vs. reference.

## Discussion

First of all, the results of this present analysis confirm that, in a large sample with a long follow-up, fasting TG is an independent risk factor for CV events. Adjusting for factors associated with both CV events and TG such as diabetes, BMI, alcohol use, HDL and non -HDL cholesterol attenuated but did not fully account for the association between TG and CV events. This is consistent with a series of studies showing a positive association of incident CV events with TG both fasting and non-fasting, in particular three large meta-analyses provided adjusted odds ratio values ranging from 1.57 (95% CI 1.10–2.24) to 1.80 (95% CI 1.49–2.19) when individuals with TG in the top tertile were compared to those with TG in the bottom tertile [26–29]. Unlike our study, a number of studies have found that the association between TG and CV risk is attenuated once adjusted for other lipid parameters, including HDL-C and non-HDL-C. An analysis conducted by the Emerging Risk Factors Collaboration (n=302,430) demonstrated that the HR for CHD as a result of elevated TG was 1.37 (95% CI 1.31– 1.42) when adjusted for non–lipid factors and became nonsignificant (0.99, 95% CI 0.94–1.05) when adjusted for HDL cholesterol and non–HDL cholesterol (0.99, 95% CI 0.94–1.05) [12]. Elevated TG levels are closely associated with higher levels of non-HDL cholesterol and low levels of HDL cholesterol [30] and this may explain why this association is weakened after adjustment for these parameters.

No significant interaction was found according to gender. This finding suggests that the risk associated with increasing TG was similar in men and women, excluding the need to identify different cut-offs of TG by gender for a better risk prediction. Other observational data have demonstrated that TG are more strongly associated with CVD risk in women than men. However, importantly, in both men and women, increasing TG were associated with increased CVD risk even among those with TG well below 150 mg/dL [31].

In the last 25 years, other Authors have demonstrated a lower cut-point for TG (100 mg/dl) to be associated with increased risk of primary and secondary CV events [32–34]. In the present study, the prognostic cut-off value of TG able to identify the subjects at risk of developing CV events was 89 mg/dL in the whole cohort: being over the cut-off, significantly led to HR>1 of developing a CV event. Therefore, our findings expand the role of TG in predicting cardiovascular risk. Guidelines acknowledge that a fasting TG level of <150 mg/dL is desirable [4,35]. Indeed, the AHA/ACC/multisociety cholesterol guideline recommended the use of elevated TG as a “risk-enhancing factor” in primary CV prevention and recommended optimizing diet and lifestyle as the first step. Emphasis on weight loss (5%-10% reduction in body weight) through healthy diet and physical activity (at least moderate 150 minutes/week or vigorous 75 minutes/week) can substantially lower TG levels by 20% to 50%. A healthy dietary pattern includes lean protein, fish, fresh fruits and vegetables, legumes, avoidance of refined foods with high glycemic index and added sugars and restricting alcohol intake [36]. If elevated TG or non-HDL-C levels remain following aggressive lifestyle intervention and statin therapy, guidelines recommend the use of TG-lowering agents [4,35]. The JELIS (Japan EPA Lipid Intervention Study) trial and the REDUCE-IT (Reduction of Cardiovascular Events with Icosapent Ethyl–Intervention Trial) trial used icosapent ethyl, a prescription grade purified eicosapentaenoic acid (EPA) [37–38]. The JELIS trial looked at more than 18,000 individuals and showed that EPA reduced major coronary events by 19% compared with the control group [37]. The REDUCE-IT trial included more than 8000 patients with CVD or diabetes with additional CV risk factors [38]. All participants were on statins with LDLs below 100 mg/dL and TG between 135 and 499 mg/dL (with a mean triglyceride level of 216 mg/dL). The patients were randomly assigned to icosapent ethyl 4 g daily or mineral oil. The composite cardiovascular endpoint was reduced by 25% over approximately 5 years, with a number needed to treat of 21.

The main strength of the study described here is to have determined on a large Italian nationwide database with a long-lasting follow-up a clear prognostic cut-off of TG, identified by the ROC curves methods and validated in multivariate models, able to identify subjects at higher CV risk. Further, if a test is used for the purpose of screening in an epidemiological context, then a cut-off value with a higher sensitivity and negative predictive value must be considered [39]. The limitations are represented by the fact that data are partially derived from a selected sample of patients referred by general practitioners to specialized centres, an underestimation of morbid events is quite likely as in other cohort studies, the design was fit to demonstrate an association but not a causality in the relationship between TG and CV events, and the analysis was based on a single TG measurement without taking into consideration the dilution bias. A recent paper based on 15,792 study participants from the Atherosclerosis Risk in Communities and Framingham Offspring studies, using fasting TG measurements across multiple exams over time, showed that the average of several TG readings provided incremental improvements for the prediction of CVD relative to a single TG measurement [31]. However, regardless of the method of measurement, higher TG were associated with increased CVD risk, even at levels previously considered “optimal” (<150mg/dL).

On the other hand, the collected database represents the largest number of Italian cases ever collected, and to our knowledge there is no one more representative of the Italian situation. Furthermore, the present analysis was limited to Italian people and its results cannot be directly applied to other ethnicities. Consequently, further studies are needed to confirm that the thresholds of TG emerging from our analyses are valid also in general populations and in other ethnicities.

**In conclusion**, the Working Group on Uric Acid and Cardiovascular Risk of the Italian Society of Hypertension confirmed that, after adjusting for potential confounders, a lower than conventional prognostic cut-off of TG able to separate subjects at risk of developing CV events can be identified in an Italian cohort.

## Sources of Funding

This work has been conducted with an unrestricted grant from the Fondazione of the Italian Society of Hypertension (grant: MIOL).

## Disclosures

Borghi has received research grant support from Menarini Corporate and Novartis Pharma; has served as a consultant for Novartis Pharma, Alfasigma, Grunenthal, Menarini Corporate, and Laboratoires Servier; and received lecturing fees from Laboratoires Servier, Takeda, Astellas, Teijin, Novartis Pharma, Berlin Chemie, and Sanofi. The authors declare no competing interests. The remaining authors have no disclosures to report.

## Data Availability

The data that support the findings of this study are available on request from the corresponding author [VT].

## Non-standard Abbreviations and Acronyms

CI: confidence interval(s)
CHD: coronary heart disease
CV: cardiovascular
HDL: high density lipoprotein
HR: hazard ratio(s)
ICD-10: International Classification of Diseases – 10^th^ Revision
LDL: low density lipoprotein
ROC: receiver operating characteristic
TG: total plasma triglycerides
URRAH: URic acid Right for heArt Health

---

**Table 1s.**
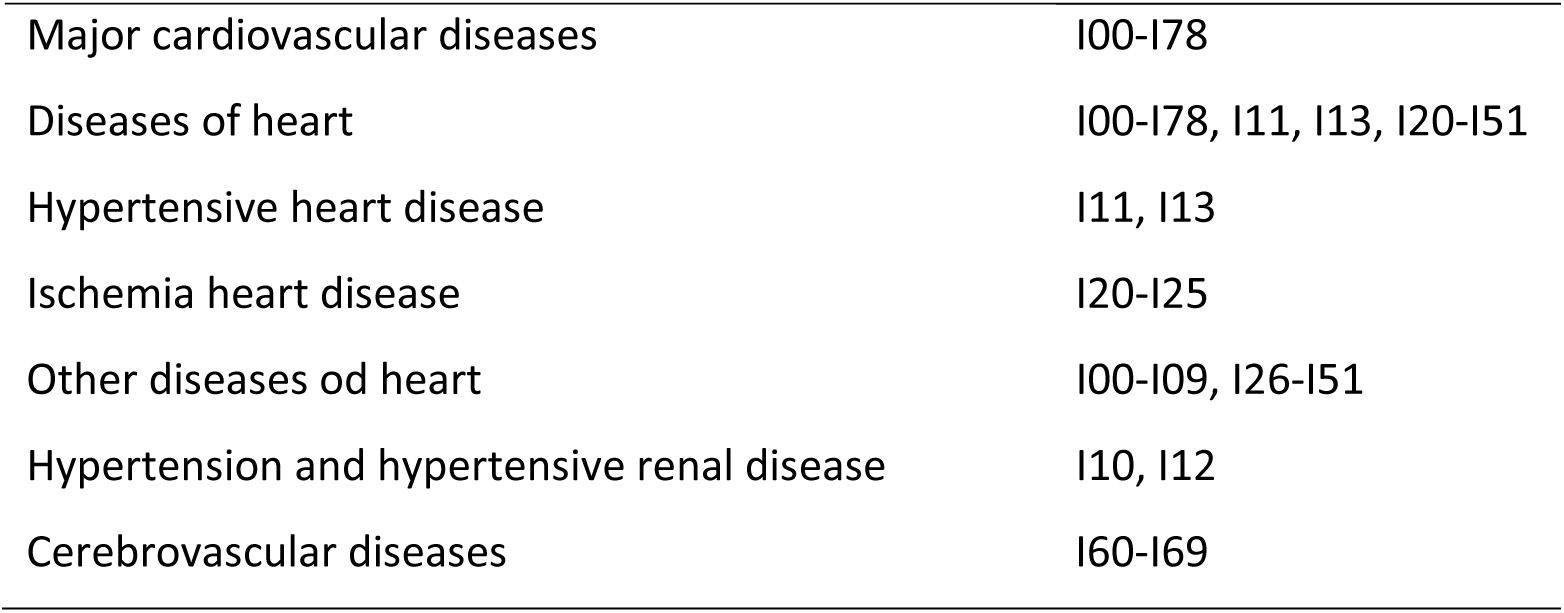
ICD10 codes used for diagnosis of cardiovascular events.

## Notes

### Competing Interest Statement

Arca has received research grant support from Aegerion, Amgen, IONIS, Akcea Therapeutics, Pfizer, Regeneron, and Sanofi; has served as a consultant for Amgen, Aegerion, Akcea Therapeutics, Regeneron, Sanofi, and Alfasigma; and received lecturing fees from Amgen, Aegerion, Merck, Pfizer, Sanofi, and Alfasigma. Borghi has received research grant support from Menarini Corporate and Novartis Pharma; has served as a consultant for Novartis Pharma, Alfasigma, Grunenthal, Menarini Corporate, and Laboratoires Servier; and received lecturing fees from Laboratoires Servier, Takeda, Astellas, Teijin, Novartis Pharma, Berlin Chemie, and Sanofi. Pontremoli has served as a consultant for and has received lecturing fees from Novartis, MSD, AstraZeneca, Boehringer-Ingelheim, Lilly, Teijin, Astellas, and Alfasigma. Desideri has received research grant support from AstraZeneca and Menarini; has served as a consultant for Servier, Menarini, FIRMA, and Sigma-Tau; and received lecturing fees from Servier, Bayer, Guidotti, Bristol Myers Squibb, DOC, and Sigma-Tau. Temporelli has received lecturing and consulting fees from Alfasigma; consulting fees from Bayer; lecturing fees from Menarini, MSD, and Servier. D'Erasmo has received research grant support from Aegerion, Amryt, Akcea Therapeutics.

### Author Declarations

Ethical Committee of the Coordinating Center at the Division of Internal Medicine of the University of Bologna (No. 77/2018/Oss/AOUBo).

